# Immunohistochemical profiles of dermatofibroma and dermatofibrosarcoma protuberans: A scoping review

**DOI:** 10.1101/2024.10.23.24316006

**Authors:** Céline S. C. Hardy, Ali Razavi, Natalie Nunez, William Fitzmaurice, Loma Dave, Amy R. Slutzky, Kaitlin Vanderbeck, Ramsay Farah

## Abstract

This review summarizes the immunohistochemical profiles of dermatofibroma (DF) and dermatofibrosarcoma protuberans (DFSP) as reported by primary literature in the past 10 years. 63 studies were included in the review, with staining information for a total of 99 unique IHC markers reported. The most commonly reported stains were CD34, SMA, S100, and FXIIIa. Most studies applied IHC either to explore descriptive features of DF or DFSP or to determine their utility in diagnostic identification of the lesions. A number of studies applied novel biomarkers which may hold promise for distinguishing DF and DFSP, namely WT1, Cx43, LSP-1, and PHH3, which demonstrated considerable expression differences between the two lesions. This review highlights the need for validation of existing and emerging IHC markers for the diagnosis of DF and DFSP.

## 1. INTRODUCTION

Dermatofibroma (DF) and dermatofibrosarcoma protuberans (DFSP) are spindle cell lesions with overlapping clinical and histologic features. DF, also known as benign fibrous histiocytoma (BFH), are benign soft tissue tumors with good prognosis. Contrastingly, DFSP are locally aggressive tumors which frequently recur. Additionally, DFSPs may undergo fibrosarcomatous transformation, which portends a higher rate recurrence and increased risk of metastasis^1,2^. Appropriate identification of DFs and DFSPs from each other and their histologic mimics is of critical importance to ensure proper clinical management. Despite their clinical divergence, these two lesions share many histologic and morphologic features. Morphologically, both are spindled lesions which may show increased cellularity and mixed fascicular, whorled or storiform growth patterns. Given their clinical and histologic similarities, it may be challenging to differentiate these two entities on histologic grounds alone, particularly in limited biopsies. Therefore, immunohistochemistry (IHC) is often used to differentiate DF from DFSP and is a mainstay in their diagnosis. Key immunohistochemical markers classically include cluster of differentiation 34 (CD34) and Factor XIIIa (FXIIIa), where CD34 is known to be strongly positive in over 90% of DFSP^3,4^, while DFs are known to be negative for CD34 but positive for factor XIIIa ^5,6^. Additional markers used to distinguish between these lesions from each other and their mimics include smooth muscle actin (SMA), S-100, and Desmin. Within both DF and DFSP, IHC can also inform or validate the presence of histologic subtypes, pathogenic/driving gene fusions, and specific pathogenic mechanisms within these lesions.

However, despite the elaborate use of IHC in classifying and studying DF and DFSP, the diagnostic specificity of the currently employed markers is imperfect. Although a number of novel markers have been suggested, constantly emerging literature coupled with a lack of validated data can lead to confusion about which markers are the most effective. For this reason, we conducted a scoping review of the literature in order to systematically report IHC staining data on these lesions, summarize the utility of existing markers, and highlight emerging markers specifically useful for distinguishing DF and DFSP. An effective summary of the current literature on the IHC staining profiles of these lesions is useful for consolidating information and providing insight into prospective biomarkers which require additional validation.

## 2. METHODS

### 2.1 Study design

This review was conducted and reported in accordance with the Preferred Reporting Items for Systematic Reviews and Meta-Analyses Extension for Scoping Reviews (PRISMA-ScR) guidelines^7^. An a priori study protocol was registered in the Open Science Framework (https://doi.org/10.17605/OSF.IO/92EWY).

### 2.2 Information sources and search strategy

Four databases – PubMed, Embase (Elsevier), Scopus (Elsevier), and the Cochrane Library – were searched for relevant articles. For each database, key concepts were defined using combinations of controlled vocabulary (where available) and keywords, to maximize search sensitivity. The PubMed search strategy is given below; full search strategies are given in Appendix X.

PubMed search strategy:

(“Histiocytoma, Benign Fibrous”[Mesh] OR “Dermatofibrosarcoma”[Mesh] OR “histiocytoma*”[tiab] OR dermatofibro*[tiab] OR DFSP[tiab] OR “nodular subepidermal fibrosis”[tiab])

AND

(“Immunohistochemistry”[Mesh] OR immunohistochem*[tiab] OR IHC[tiab] OR immunostain*[tiab]) Database search strategies limited search results to publication dates over the past 20 years (2003-present). Final searches for all databases were run on June 24, 2024.

### 2.3 Eligibility criteria

Studies were selected for inclusion in the review if they:

- evaluated protein expression by immunohistochemistry (IHC), specifically immunoperoxidase staining, of tissue specimens from human subjects of histologically confirmed cases of cutaneous DF (including benign fibrous histiocytoma, cutaneous fibrous histiocytoma, and epithelioid fibrous histiocytoma) or DFSP;
- reported the number of lesions evaluated and the fraction of lesions positive or negative for a given marker for greater than 5 lesions;
- focused on evaluating the tumor cell compartment (as opposed to stromal or immune cell profiling); and
- were published in English, French, or Spanish.

The following types of studies were excluded from the review: review articles, case reports, conference abstracts, in vitro studies, and animal studies.

### 2.4 Article screening

Results from all database searches were uploaded to Covidence (Veritas Health Innovation, Melbourne, Australia). Duplicate records were identified and removed automatically by Covidence, with all duplicate pairs manually confirmed (AS). Then two reviewers (AR, WF, NN, and/or LD) independently screened each title/abstract for inclusion in the review, with any conflicts resolved via discussion with content experts (RF and/or KV). After title/abstract screening was complete, a decision was made to limit the review to studies published over the past 10 years (2013-present), and to exclude case studies and case series evaluating fewer than five lesions. These decisions were made both due to practical/time constraints and in consideration of the relevance of IHC markers evaluated, pertinence and consistency of antibody clones and staining protocols used. Full-text articles were then screened independently by two reviewers (AR, WF, NN, and/or CH), with any conflicts resolved as described above.

### 2.5 Data abstraction

Data collection was performed first by one of two independent reviewers (CH, AR) and then independently verified by the second reviewer. The following data were collected and charted using Microsoft Excel:

- Study characteristics (article title, publication date, study aim, study geographic location)
- Lesion information (type and number of lesions studied, sample/specimen type, histologic information if available)
- IHC characteristics (IHC stains performed, method of IHC evaluation, specific antibodies used, staining expression results for lesions of interest)

## 3. RESULTS

### 3.1 Selection and characteristics of sources of evidence

Database searches identified 2506 distinct articles. Following title/abstract and full text screening, 63 articles were selected for inclusion in the review^8–70^ (**Fig. 1**). Of those 63 studies, 15 contained IHC data for DF only, 28 for DFSP only, and 20 studies contained data for both DF and DFSP. Complete characteristics of data extracted from all articles -- including geographic location, date of sample collection, and study aim -- are summarized in supplemental **Table S1**.

**Figure 1.**
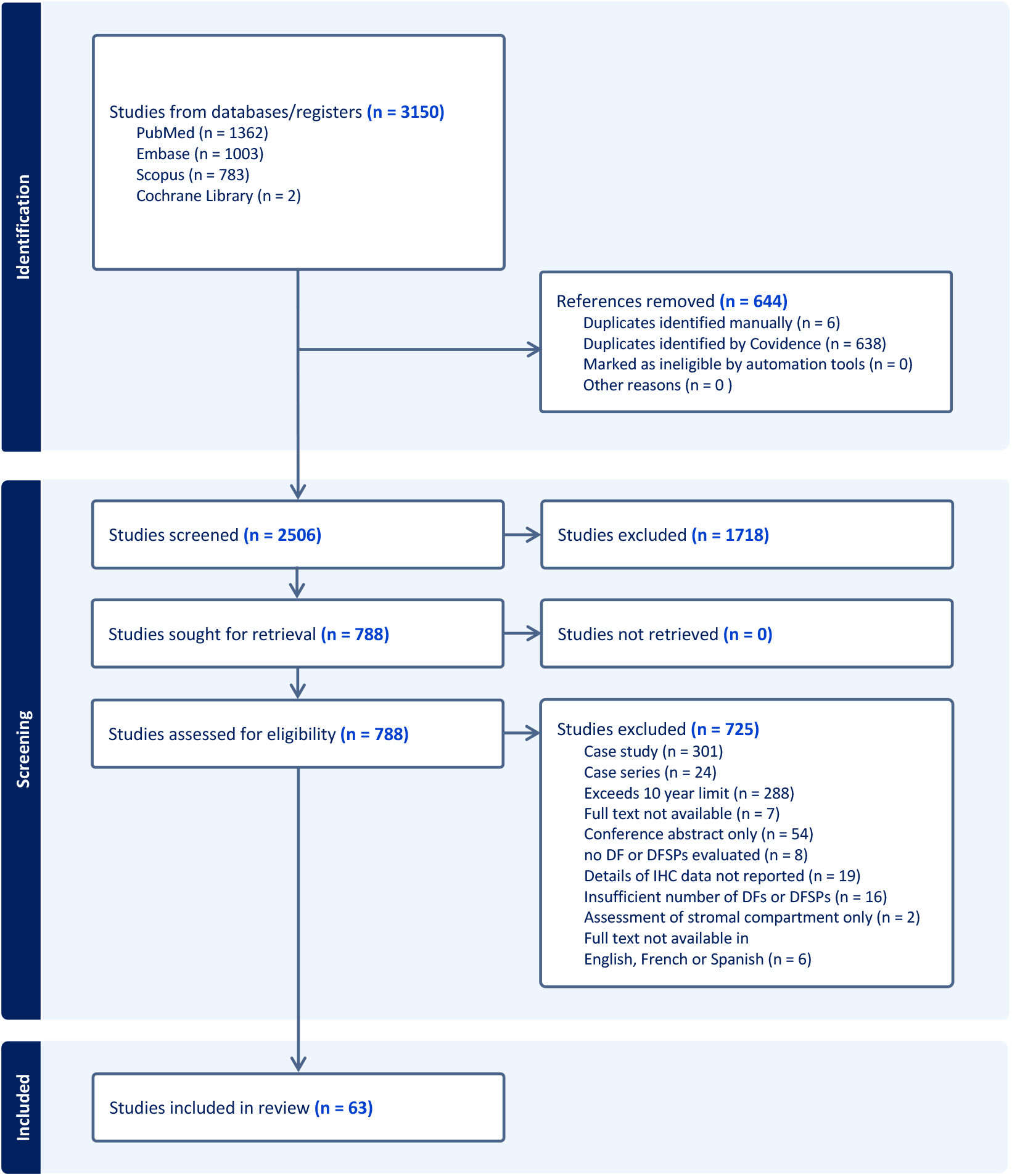
PRISMA flow diagram indicating the article selection process.

### 3.2 Immunohistochemical markers applied in DF and DFSP

Across all studies, a total of 99 IHC markers were evaluated, of which the most discussed markers included CD34, SMA, S100, FXIIIa, Desmin, and Ki-67 (**Fig. 2**). Cluster of differentiation 68 (CD68), anaplastic lymphoma kinase (ALK), epithelial membrane antigen (EMA), and pan-cytokeratin (PanCK) were also frequently discussed (**Fig. 2**). The purpose of IHC application in these studies could be broadly categorized as descriptive, diagnostic, or prognostic. Summary tables of IHC biomarkers applied and their indication or purpose of application in DF and DFSP are summarized in **Table 1** and **Table 2**, respectively. The majority of IHC markers were applied for descriptive or diagnostic purposes. Descriptive studies focused primarily on characterizing expression of a given marker in specific entities, determining associations of expression with variant histology, or evaluating the role of expression in disease pathogenesis. Of the studies evaluating IHC staining with respect to its diagnostic utility, 37 studies assessed a role for IHC in distinguishing histologic mimics from either DF or DFSP, while 15 studies contained data focused specifically on distinguishing DF and DFSP from each other.

**Figure 2.**
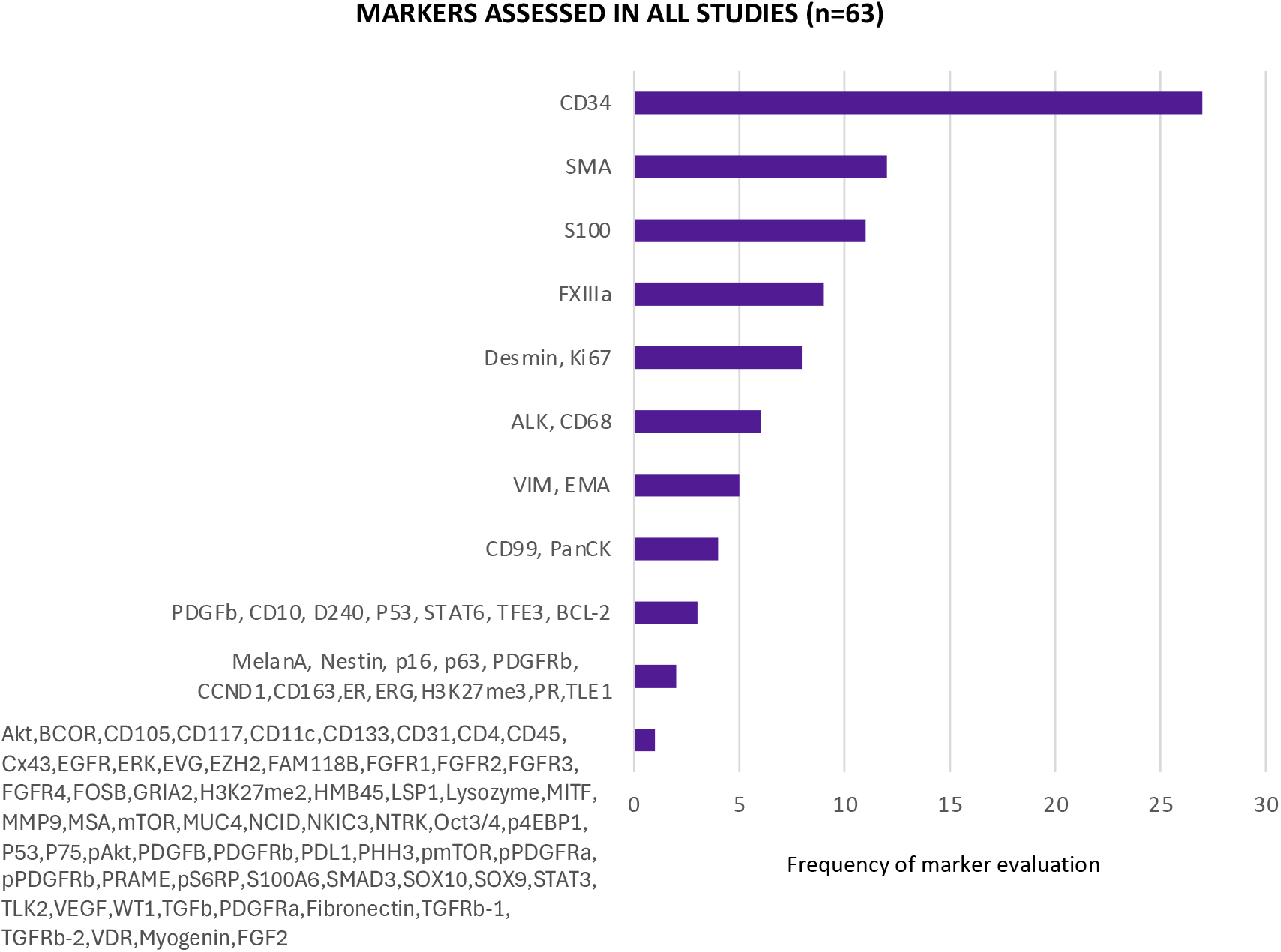
Frequency of IHC biomarker use across all studies (n=63). Bar plot indicating the frequency of IHC biomarkers assessed across all studies. Bars indicate the number of studies which evaluated a given marker.

**Table 1.** Immunohistochemical biomarkers applied in DF and their primary indication as evaluated in the studies reviewed.

| Indication | Markers | Utility/Findings |
| --- | --- | --- |
| Diagnostic | CD34, FXIIIa, S100, SMA, Desmin, Vimentin | Classical diagnostic markers |
|  | FOSB | Distinguish from pseudomyogenic hemagioendothelioma<sup>31</sup> |
|  | SOX9 | Distinguish from AFH<sup>16</sup> |
|  | TLE1, BCOR | Distinguish from AFH<sup>48</sup> |
|  | STAT6 | Distinguish from SFT<sup>20</sup> |
|  | MITF, NKI/C3 | Compare to expression in cellular neurothekoma<sup>9</sup> |
|  | TFE3 | Compare expression to desmoid type fibromatosis<sup>70</sup> and evaluate histopathologic features<sup>36,45</sup> |
|  | ERG<sup>29,67</sup>, WT1<sup>50</sup>, p75<sup>64</sup>, D2-40<sup>14,53</sup> | Distinguish from DFSP and its mimics |
|  | CD10<sup>39,60</sup>, CCND1<sup>39</sup>, LSP1<sup>34</sup>, CD68<sup>34,42,45,47</sup> | Distinguish from DFSP |
|  | CD99 | Distinguish from DFSP and describe expression<sup>37,60</sup> |
| Descriptive | MUC4 | Describe features of lipidized fibrous histiocytoma<sup>42</sup> |
|  | FGFR1-4, FGF2 | Understand expression patterns and role in pathogenesis<sup>32</sup> |
|  | Akt, mTOR, STAT3, ERK, CCND1 | Understand expression pattern and assess role in recurrence<sup>49</sup> |
|  | CD163<sup>45,55</sup>, CD11c, CD31, CD45, lysozyme, and CD4<sup>55</sup> | Characterize the immunophenotype of solitary xanthogranuloma in comparison to its differential diagnoses (including BFH) |
|  | NCID<sup>38</sup>, PRAME<sup>18</sup> | Describe expression in various fibroproliferative diseases |
|  | TGFb, TGFRb-1/2, SMAD3, VDR | Describe expression in DF, cutaneous leiomyomas, and uterine leiomyomas<sup>19</sup> |
|  | PanCK<sup>42,45</sup>, SOX10, p63, and EMA<sup>45</sup>, P16<sup>59,69</sup>, P53<sup>33,41</sup>, CD105, CD133, Oct3/4, CD117, and ALK<sup>60</sup>, PDGFRα/β, PDGFα/β<sup>46,60</sup>, Nestin<sup>46</sup> | Describe expression |
| **Prognostic** | Ki-67, MMP9 | Association with high recurrence rate<sup>11</sup> |

**Table 2.**
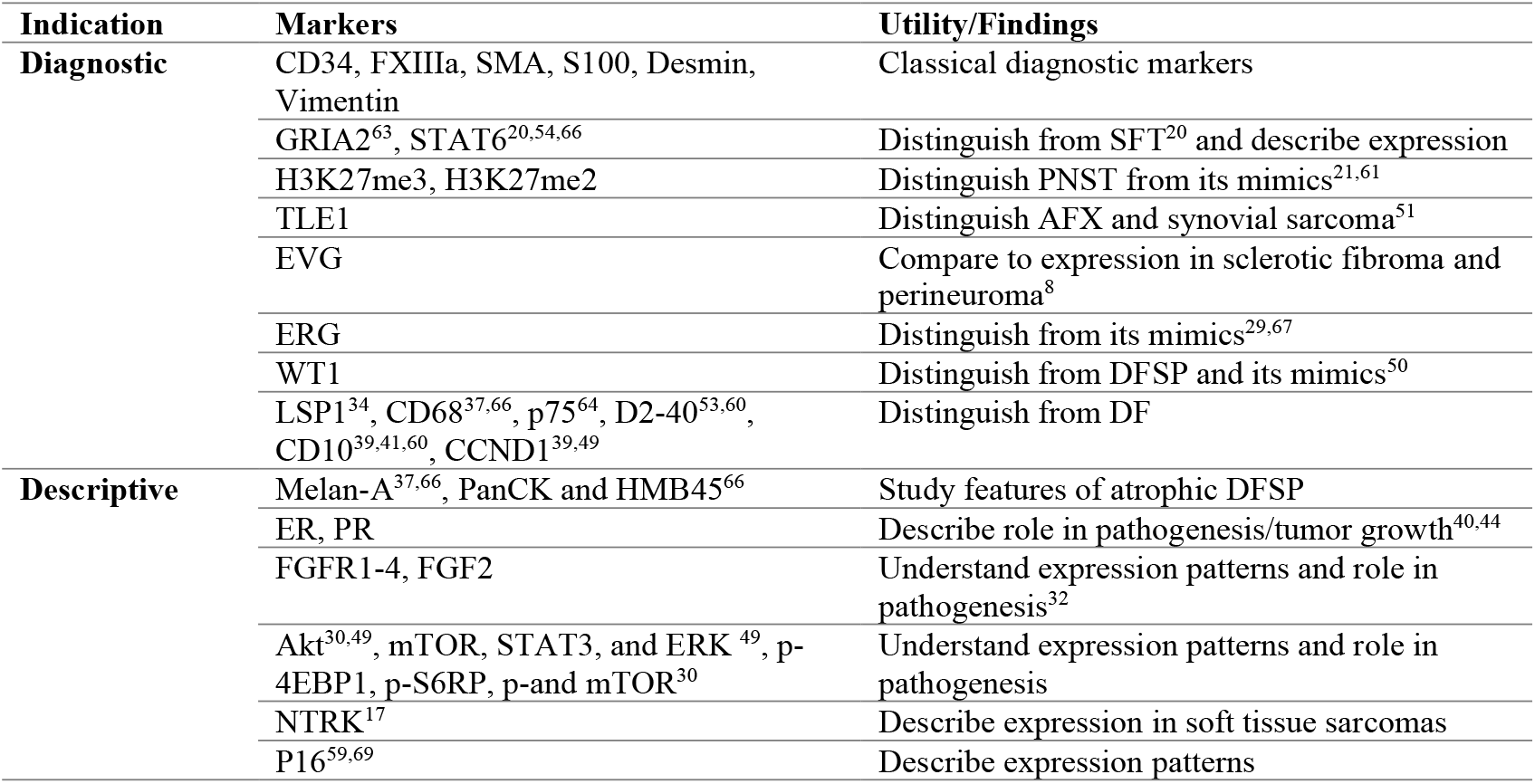
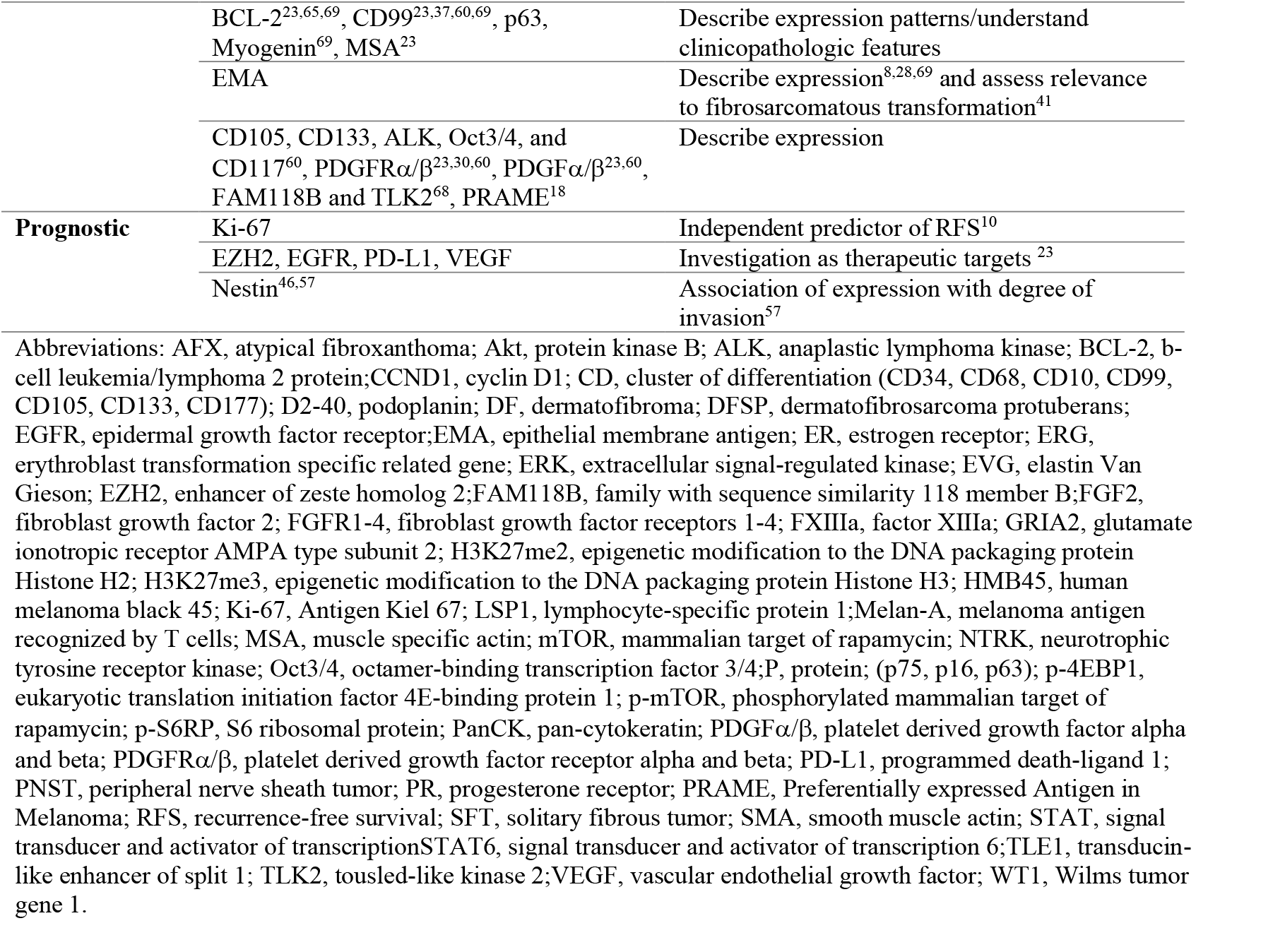
Immunohistochemical markers applied in DFSP and their primary indication as evaluated in the studies reviewed.

### 3.3 Immunohistochemical markers for distinguishing DF and DFSP

When focusing on studies aiming to characterize differences in immunohistochemical expression between DF and DFSP (n=13), a total of 30 IHC markers were evaluated. CD34 and Factor XIIIa were the most frequently employed markers for distinguishing DF and DFSP in the reviewed studies (**Fig. S1**). Summary statistics of the staining expression of classical markers in DF compared to DFSP across all studies are provided in **Table 3**. CD34 was frequently strongly positive in DFSPs (95.8%), while negative in DF (15.5%), and contrastingly Factor XIIIa was positive in DF (86.2%) but more frequently negative in DFSP (24.9%) (**Table 3**). SMA, ALK, CD99, and CD68 were more frequently positive in DF than DFSP, while S100 and Desmin tended to be negative in both (**Table 3**). Many studies also provided data regarding more novel markers evaluated for distinguishing between the two lesions, these included: Wilm’s tumor 1 (WT1), ETS transcription factor (ERG), podoplanin (D2-40), phosphohistone H3 (PHH3), p75, connexin 43 (Cx43), fibroblast growth factor 2 (FGF2), fibroblast growth factor receptor 4 (FGFR4), lymphocyte specific protein 1 (LSP-1), p16, and Ki-67 (**Table 4)**. Of these, studies evaluating WT1, Cx43 and LSP-1 demonstrated the greatest difference in expression between DF and DFSP. Complete expression data for all markers across all studies are available in supplemental **Table S2**.

**Table 3.**
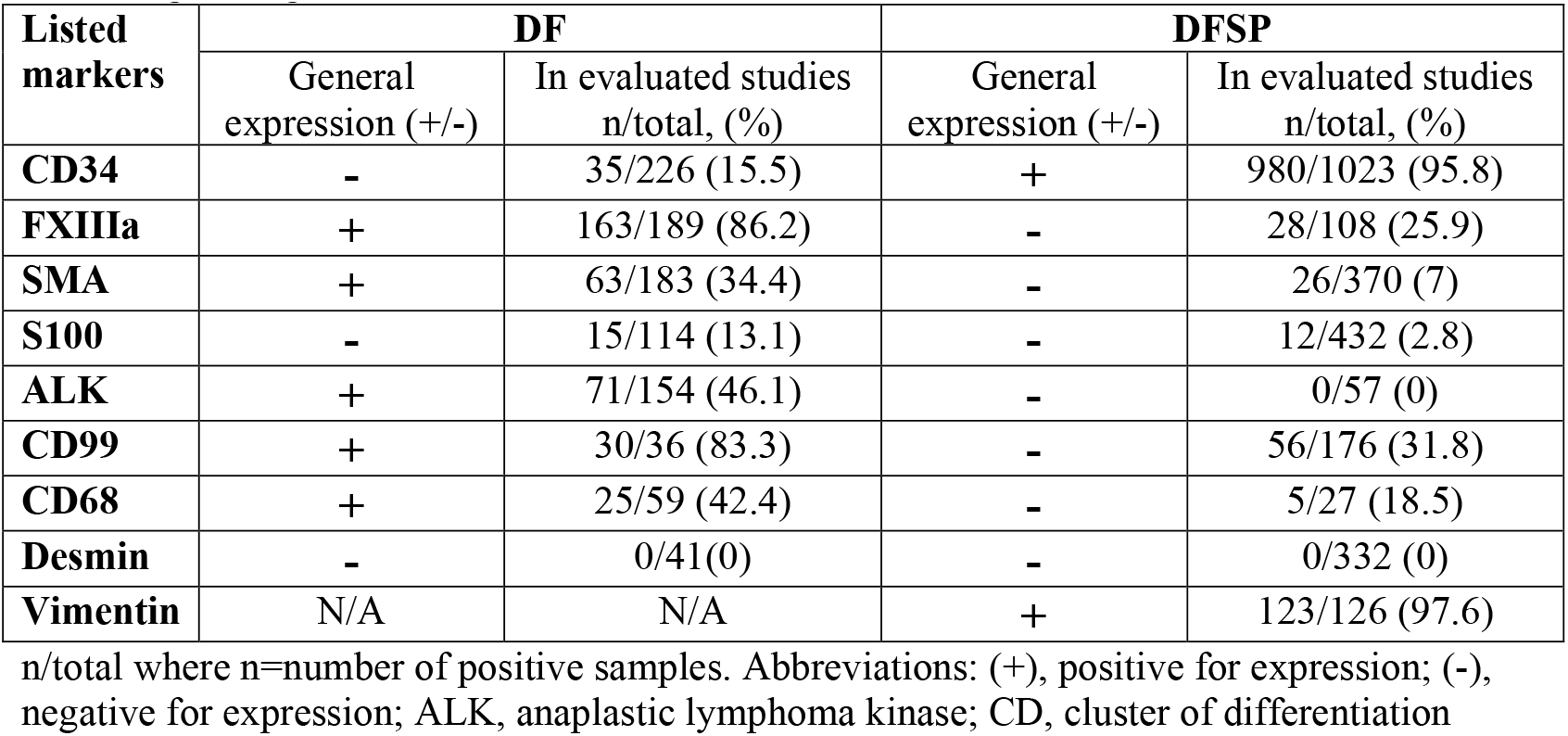
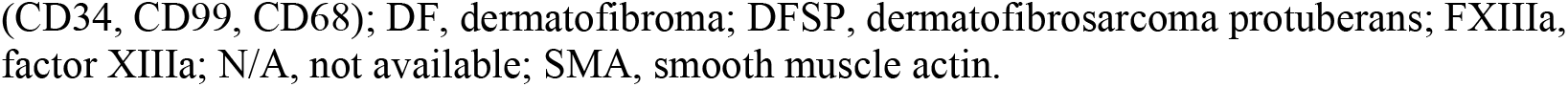
General trends of immunohistochemical expression for classical/well described markers for distinguishing DF and DFSP.

**Table 4.** General trends of immunohistochemical expression for novel/emerging markers evaluated in distinguishing DF and DFSP.

| Marker | Evidence (n positive/total) | Study |
| --- | --- | --- |
| **WT1** | Negative in DF (0/15), positive in DFSP (54/57) | Piombino et al. (2021)<sup>50</sup> |
| **Cx43** | Positive in DF (16/16), Negative in DFSP (0/13 positive) | Fernandez-flores et al. (2023)<sup>26</sup> |
| **LSP-1** | Positive in DF (18/20), negative in DFSP (2/20) | Jin et al. (2015)<sup>34</sup> |
| **ERG** | 87/97 DF positive with variable (50-89% cells) positive, 0/64 DFSP positive | Yamada et al. (2023)<sup>67</sup> |
|  | 15/15 DF positive, 6/9 DFSP positive | Hengy et al. (2022)<sup>29</sup> |
| **PHH3** | DF mean mitotic count 1.8 mm<sup>2</sup>, DFSP mean mitotic count 0.5/mm<sup>2</sup> | Agarwal et al. (2017)<sup>12</sup> |
| **Ki-67** | Mean proliferative index in DF: 39.2mm<sup>2</sup>, DFSP: 12.6 mm<sup>2</sup> | Agarwal et al. (2017)<sup>12</sup> |
| **P75** | 0/30 DF positive, 17/32 DFSP positive | West et al. (2014)<sup>64</sup> |
| **D2-40** | Positive in DF (49/70) | Aslam et al. (2021)<sup>14</sup> |
|  | Positive in DF (23/30) and negative in DFSP (2/15 positive) | Sadullahoglu 2017<sup>53</sup> |
|  | DF 4/12 positive, DFSP 2/57 positive | Song et al. (2017)<sup>60</sup> |
| **Oct3/4** | 0/12 DF, 0/57 DFSP | Song et al. (2017)<sup>60</sup> |
| **CD10** | 12/12 DF, 23/57 DFSP | Song et al. (2017)<sup>60</sup> |
|  | 13/15 DF, 15/15 DFSP | Kouki et al. (2021)<sup>39</sup> |
|  | 12/15 DF, 15/15 DFSP | Kouki et al. (2021)<sup>30</sup> |
| **FGF2 and FGFR4** | Positive in DF (20/20), variable expressivity in DFSP (4/6 positive) | Ishigami et al. (2013)<sup>32</sup> |
| **FGFR1-3** | All cases negative in both DF (0/20) and DFSP (0/6) | Ishigami et al. (2013)<sup>32</sup> |
| **P16** | Positive in both DF (18/19) and DFSP (4/5) | Smith et al. (2016)<sup>59</sup> |

Abbreviations: CD, cluster of differentiation (CD10); Cx43, connexin 43; D2-40, podoplanin; DF, dermatofibroma; DFSP, dermatofibrosarcoma protuberans; ERG, erythroblast transformation specific related gene; FGF2, fibroblast growth factor 2; FGFR1-3, fibroblast growth factor receptors 1-3; FGFR4, fibroblast growth factor receptor 4; Ki-67, Antigen Kiel 67; LSP1, lymphocyte-specific protein 1; Oct3/4, octamer-binding transcription factor 3/4; P, protein (p75, p16); PHH3, phosphohistone H3; WT1, Wilms tumor gene 1.

## 4. DISCUSSION

In this study, we summarize the applications of IHC markers in DF and DFSP and their utility for distinguishing these lesions as reported in the literature over the past 10 years. In this review a total of 99 unique IHC markers were reported in the 63 studies evaluated. Interpretation of the expression data of these markers and the utility of key markers for distinguishing DF and DFSP will be elaborated upon in the following sections.

### 4.1 Accuracy of classical markers for distinguishing DF and DFSP

Classical markers for diagnosis of DF and DFSP such as CD34, FXIIIa, S-100, and Desmin all demonstrated expression in expected proportions and patterns according to the literature^71^. However, it is important to note that despite these overall trends there was marked intra-study variability for most markers. For example, even CD34, one of the most specific markers positive in DFSP, was weakly or focally positive in ∼15% of DFs^42,53,64^. Most commonly, conventional DFs did show negativity for CD34, however many DFs which contained variant histology were specifically noted to have focal CD34 expression. This highlights previous observations that application of CD34 alone is insufficient, and the need for additional markers to clarify the identity of lesions where CD34 expression is ambiguous or present focally. Interestingly, expression of classical markers CD99 and CD68 were reported of variable diagnostic utility to distinguish DF and DFSP. While one study showed that DFs have higher frequency of CD99 positivity compared to DFSPs^37^ and additional data supports there being evidently low expression of CD99 in DFSPs (0% positivity)^23,69^, data from Song et al. shows 50% positivity in both lesions^23,37,60,69^. Altogether, there remains work to be done to determine the expression in DF compared to DFSP of classical markers using additional sample sets. Beyond these diagnostic applications, there is a need to better understand the expression of classical markers within these lesion types in regions with variant histology.

### 4.2 Emerging markers for distinguishing DF and DFSP

Given the imperfect specificity of classical markers, there is an inherent need to identify and validate the utility of more novel markers for diagnostic dilemmas. A number of the reviewed studies assessed a role for novel markers in distinguishing DF and DFSP from each other or their mimics. Promising emerging IHC markers included WT1, LSP-1, Cx43, and a combined Ki67 and PHH3 score, which warrant further investigation^12,26,34,50^.Piombino et al. assessed the expression of WT1, a master transcriptional/translational regulator in cell differentiation in a series of soft tissue neoplasms of mesenchymal origin. They found that cytoplasmic WT1 expression was absent in all evaluated DFs, but positive in 94.7% of DFSPs^50^. Similar to WT1, connexin 43 (Cx43) was evaluated by Fernandez-Flores et al. because of its suspected role in pathogenesis. Specifically, with respect to propensity for invasion given that connexins are involved in mediating cell-cell adhesion. Cx43 demonstrated excellent specificity for the lesions, with 100% positivity in DF and 0% positivity in DFSP^26^. Another promising marker studied was Leukocyte specific protein 1 (LSP-1) which was also highly positive in DF, while negative in DFSP^34^.

Although Ki-67 is traditionally thought of as a prognostic, rather than diagnostic biomarker, Agarwal et al. identify potential utility of Ki-67 for distinguishing DF and DFSP in a combined scoring system with PHH3^12^. PHH3 is expressed in M phase and is therefore useful in determining mitotic counts. The goal of this study was to assess the utility of a combined scoring system incorporating both the Ki67 index and PHH3 mitotic count, where they found that the mean Ki67 proliferation index and mitotic count were significantly higher in DF compared to DFSP^12^. Interestingly, this study may highlight that the integration of multiple features or biomarkers can provide more comprehensive information than looking for one “silver bullet” IHC marker to clarify the diagnosis.

D2-40 is another emerging marker for which a number of studies suggested utility, however these data were contradictory and require additional validation. A number of studies suggest D2-40 for aiding in the differential diagnosis of DF and DFSP, where D2-40 is positive in DF and negative in DFSP^14,53^. However, data from Song et al. demonstrated only 33% positivity in DF^60^. Like D2-20, markers such as ERG, CD10, and p75 have some data to indicate utility, but require further investigation due to conflicting data, or low sample numbers. Across all markers, it should be noted that small sample size, differences in antibody clones, staining protocols, and staining evaluation methodology remain a challenge that likely contribute greatly to the observed variability across studies evaluating a given marker.

## 5. Conclusions

In this review, we report the IHC expression in DF and DFSP of traditional diagnostic markers as well as novel markers which may hold diagnostic utility according to the literature from the past 10 years. Of the markers discussed with diagnostic relevance, traditional markers such as CD34, SMA, and FXIIIa were the most common and demonstrated expected expression patterns in DF and DFSP. Additionally, studies investigated novel markers for distinguishing DF and DFSP such as WT1, Cx43, LSP-1, and PHH3/Ki-67. Future studies should seek to continue investigating the expression of these markers and their potential utility for diagnostic application.

## Acknowledgements

the authors have no acknowledgements to report.

## Data availability

Data supporting the findings of this study are available within the article and/or its supplementary material.

## SUPPLEMENTAL MATERIAL

**Table S1**. Study characteristics data extracted from the included studies (n=63).

**Table S2**. IHC expression data extracted from the included studies (n=63).

**Figure S1. Frequency of IHC biomarker use in studies distinguishing between DF and DFSP (n=15)**. Bar plot indicating the frequency of IHC biomarkers assessed across studies which aimed to differentiate DF and DFSP. Bars indicate the number of studies which evaluated a given marker.

