## Supplementary figures and images for "Immunohistochemical profiles of dermatofibroma and dermatofibrosarcoma protuberans: A scoping review"

### Supplemental figure 1

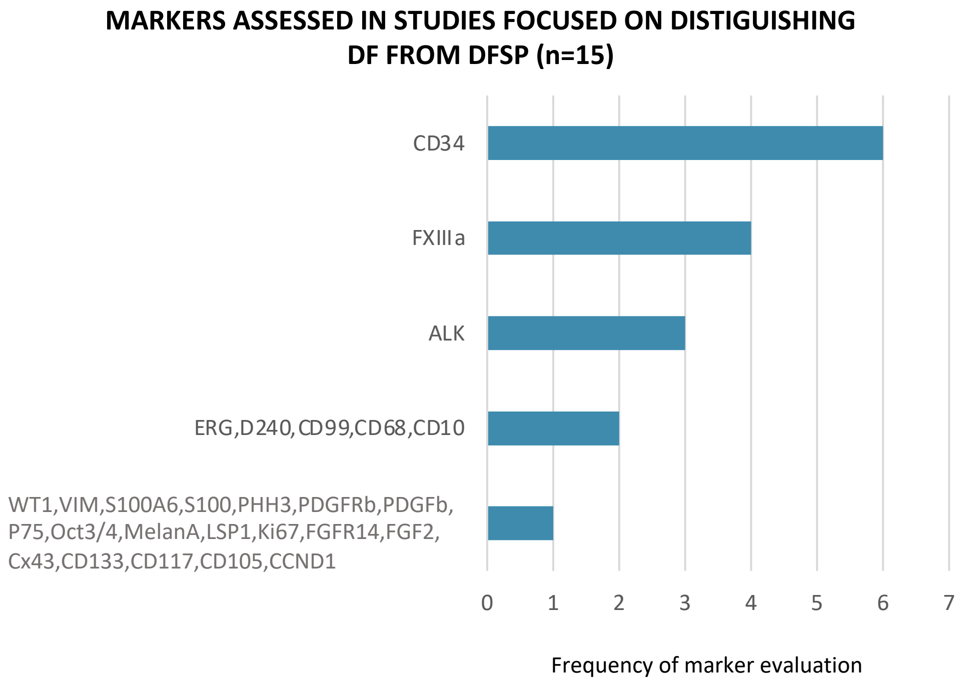
